# IS ESPORTS ASSOCIATED WITH INCREASED HEALTH RISKS? A CROSS-SECTIONAL COMPARISON OF MUSCULOSKELETAL PAIN PREVALENCE AMONG YOUNG DANISH ESPORTS PLAYERS AND HANDBALL PLAYERS

**DOI:** 10.1101/2022.09.19.22279922

**Authors:** Frederik Sand Hansen, Mathias Lyngs, Mathias Dyg Hyllested Lauridsen, Christian Lund Straszek

**Affiliations:** Department of Physiotherapy, University College of Northern Denmark (UCN), Aalborg, Denmark; Center for General Practice at Aalborg University, Aalborg, Denmark; Department of Health Science and Technology, Aalborg University, Aalborg, Denmark

**Author notes:** **Correspondence:** Christian Lund Straszek, Address: Selma Lagerløfs Vej 2, 9220 Aalborg East, Denmark.

## Abstract

**Background:** There is a high prevalence of musculoskeletal (MSK) pain and extensive training volume among professional and amateur esport players. MSK pain has been found to limit esports participation in 6% of players. Further, there is an increasing concern that the training volume may lead to activity-limiting burnout in esports. However, it remains to be investigated if MSK pain and activity-limiting burnout is more prevalent in esports compared to traditional sports such as handball. The objectives were 1) to compare MSK pain prevalence between esports players and handball players and 2) to investigate if MSK pain is associated with training volume in the two groups.

**Methods:** Eligible players had to engage in either structured esports or handball and be 15-25 years of age to participate in this questionnaire-based cross-sectional study. Esports players had to engage in esports primarily through a computer-based game. Demographic data, self-reported MSK pain prevalence, training volume, sleep patterns, physical activity level and activity-limiting pain and burnout were obtained through online questionnaires. The primary outcome was any MSK pain during the previous week (yes/no).

**Results:** 76 esports players and 175 handball players were included. 48% of esports players and 80% of handball players experienced MSK pain during the previous week. The likelihood of MSK pain was significantly lower in esport compared to handball (OR 0.24, 95%CI [0.13-0.43], Chi^2^ p-value < 0.001). No significant differences in training volume among participants with or without MSK pain were found in neither esports (p-value = 0.727) nor in handball (p-value = 0.128). There was a significant difference in training volume with esports player practicing for additional 12 hours per week compared to handball players (p-value < 0.001). The occurrence of activity-limiting burnout was high in both esports (34%) and handball (37%).

**Conclusion:** These findings suggest that young esports players are not at increased health risk in terms of experiencing MSK pain compared to young players participating in handball. This despite esports players practiced their activity for 12 hours more per week compared to handball players. More than 1 in every 3 had experienced activity-limiting burnout in both esports and handball despite a significant difference in weekly training volume. This indicate that other factors besides type of activity and training volume may influence the occurrence of activity-limiting burnout.

## BACKGROUND

Competitive gaming (esports) has gained increasing popularity among adolescents and young adults (1,2). Esports players compete in various digital games through a computer, console, or smartphone both online and at large events with thousands of spectators and millions of dollars in prize money (3-5). With the increased recognition and commercial interest in esports, more awareness has been directed toward esports players’ mental and physical health. As such, larger esports organizations have employed performance teams consisting of health professionals such as mental coaches, physiotherapists, and medical doctors to help the players stay in good health and ready for competition. In relation, musculoskeletal (MSK) pain, especially related to the back and wrist, was found to be highly prevalent among professional esports players (5,6). Moreover, one study found that 42% of young amateur esport players experienced (MSK) pain (1). This is important as amateur players may not have the same access to healthcare as professional players do. Although continuously debated, one of the underlying mechanisms for MSK pain in esports is thought to be the high training volume to which many esports players are exposed.

The average training volume among amateur esports players was found to be more than 24 hours/week. One study showed that esports players with MSK pain during the previous week had a significantly lower training volume than those without MSK pain (1). As such, MSK pain may limit participation among amateur esports players. Notably, the weekly training volume among professional esports players could be three times higher than amateur players (6). This is important as an emerging concern in esports is related to mental health issues and activity-limiting burnout, which may be associated with the high training volume (7). As such, health professionals have raised several concerns about esports and have called for additional research on health in esports (4). As esports appeal to the younger population, additional knowledge on health and risk factors is warranted to support the young players.

Although the prevalence of MSK pain is high in esports, it remains unknown if the risk of MSK pain is higher in amateur esports compared to that of other activities that adolescents participate in. In Denmark, handball is the second most popular sport among adolescents and young adults only surpassed by football (8). As handball and esports appeal to the same age group, it is worth investigating if the risk of MSK pain is higher in esports compared to handball. In addition, it remains to be investigated if MSK pain is continuously associated with decreased training volume across different populations of amateur esports players, thus replicating the results from Lindberg et al 2020 (1). Lastly, it is currently unknown if activity-limiting burnout is more common in esports compared to other activities such as handball.

Therefore, the aims of this study were 1) to compare MSK pain prevalence among amateur esports players and handball players and 2) to investigate if MSK pain is associated with training volume in esports and handball. We hypothesised that MSK pain prevalence would be higher among handball players and that MSK pain would decrease training volume in both esports and handball.

## METHOD

This questionnaire-based cross-sectional study was conducted at the Department of Physiotherapy at University College of Northern Denmark, Aalborg, Denmark. The study protocol was developed from the Strengthening the Reporting of Observational Studies in Epidemiology (STROBE) statement (9). The players received written study information before providing written informed consent to participate. Approval to conduct the study was sought from the Ethical Committee of Northern Denmark. The committee replied that no approval was necessary for the current study (journal number: 2022-000764).

### Eligibility Criteria

To reach a representative study sample, we aimed to include 150 players from both esports and handball. Eligible players had to be between 15 and 25 years of age and participate in structured esports or handball (defined as training with a coach present). Esports players were required to primarily participate in esports through a computer-based game. As such, eligible esports players and handball players were sought out at community-based teams, professional organizations, and educational institutions via emails, phone calls and through the authors’ network in esports and handball. Players were in addition recruited from competitive tournaments and through social media.

### Distribution of questionnaire

The questionnaire was developed and distributed through SurveyXact. The questionnaire was pilot tested among both esports players and handball players to evaluate completion time and relevance.

### Player characteristics

All players were asked to provide name, email, phone number, age, sex, height, weight, level of play (amateur, semi-professional or professional) and to state in which context they participated in their activity (e.g., professional team or educational institution). Esports players were required to state the title of their primary competitive game.

### Outcome measures

#### Primary outcome

The primary outcome was ‘Have you experienced pain in your body during the previous week?’. This question was answered with ‘yes’ or ‘no’. This method was used as the primary outcome in a previous study investigating MSK pain prevalence in amateur esport players (1).

#### Pain measures

Participants who answered ‘yes’ to the initial question of MSK pain were subsequently asked to state their primary pain site. Pain intensity was assessed as worst pain during the previous week at the primary pain site with a 11-point Numeric Pain Rating Scale (0 = no pain, 10 = worst possible pain). Pain frequency was investigated with a 5-point rank scale ranging from daily to seldom. The prevalence of activity-limiting pain was assessed with a single question “Have you had difficulties participating in your sports due to your pain?” which could be answered with ‘yes’ or ‘no’. These pain measures were used in previous research to assess activity-limiting pain among adolescents with MSK pain who participated in sports and esports (1,10).

#### Activity-limiting burnout

Activity-limiting burnout among esports players and handball players was investigated with a single question, ‘Have you within the previous 3 months experienced the feeling of burnout to such an extent that you could not participate in your sports?’ This question was answered with ‘yes’ or ‘no’.

#### Training volume

Training volume was assessed with three individual questions. Firstly, structured training volume was defined as the number of hours per week each player practised esports or handball with a coach present. Secondly, the number of hours per week each player practised esport or handball without a coach present was defined as unstructured training volume. Lastly, the number of hours per week each player was engaged in competition or tournament activities was assessed. In the current study, the total weekly training volume was defined as the sum of weekly structured and unstructured hours of training in esport and handball. This strategy was used in a previous study to investigate training volume in esports (1). Using this approach, it was possible to compare the findings from the current study with previous findings.

#### Physical activity levels

Physical activity levels were assessed with two separate questions. The first question assessed the number of days per week each player was physically active at moderate intensity. The second question investigated the number of days per week each player was physically active at high intensity. Each question could be answered on an 8-point rank scale from ranging from 0 to 7. These two questions were supplemented with the following text: ‘*Moderate intensity is physical activity where you feel shortness of breath, but you can keep a conversation going. High intensity is physical activity where you feel shortness of breath and you experience difficulties keeping a conversation going*’. These questions were framed based on the Danish national recommendations for physical activity among children, adolescents, and young adults (11).

#### Use of analgesics

The frequency of analgesic use due to pain in the previous 3 months was assessed among players who had had indicated analgesic use with a 5-point rank scale ranging from daily use to seldom use.

#### Sleep patterns

Sleep patterns were assessed as the quantity of sleep and sleep quality. Sleep quantity was investigated as the average hours of sleep each player would get during the night. Sleep quality was assessed with four separate questions which were: 1) trouble falling asleep in the evening, 2) waking up several times during the night, 3) trouble sleeping through the night and, 4) feeling tired in the morning when waking up. Each of these four questions was answered on a 4-point rank scale (1: No, not at all, 2: Yes, some nights/mornings, 3: Yes, most nights/mornings, 4: Don’t know). Similar questions were used in previous studies to assess sleep patterns in adolescents and young adults (1,10).

### Data analysis

All statistical analyses were undertaken in IBM SPSS version 28. The Chi2 test was used to compare MSK pain prevalence between the two activities, and the Odds Ratio was used to express the association between type of activity and MSK pain. To investigate whether the training volume differed between players with and without MSK pain among players within the two groups, we used the independent sample t-test to compare total weekly training volume (total hours of structured and unstructured training per week). A p-value <0.05 was considered statistically significant. Data from two handball players were missing for measures related to analgesic use, pain frequency, pain location, pain intensity and activity-limiting pain. Further, data from four handball players were missing for measures related to physical activity levels, activity-limiting burnout, and sleep patterns.

## RESULTS

From April 20^th^ 2022 to July 6^th^ 2022, 279 players responded to the questionnaire. Of these, 19 did not respond to the primary outcome, and 9 did not meet the eligibility criteria. We included 76 esports players and 175 handball players (251 in total) from Denmark. See Table 1 for additional data regarding player characteristics and Table 2 for findings on the use of analgesics, physical activity level, activity-limiting burnout, and sleep patterns.

**Table 1.**
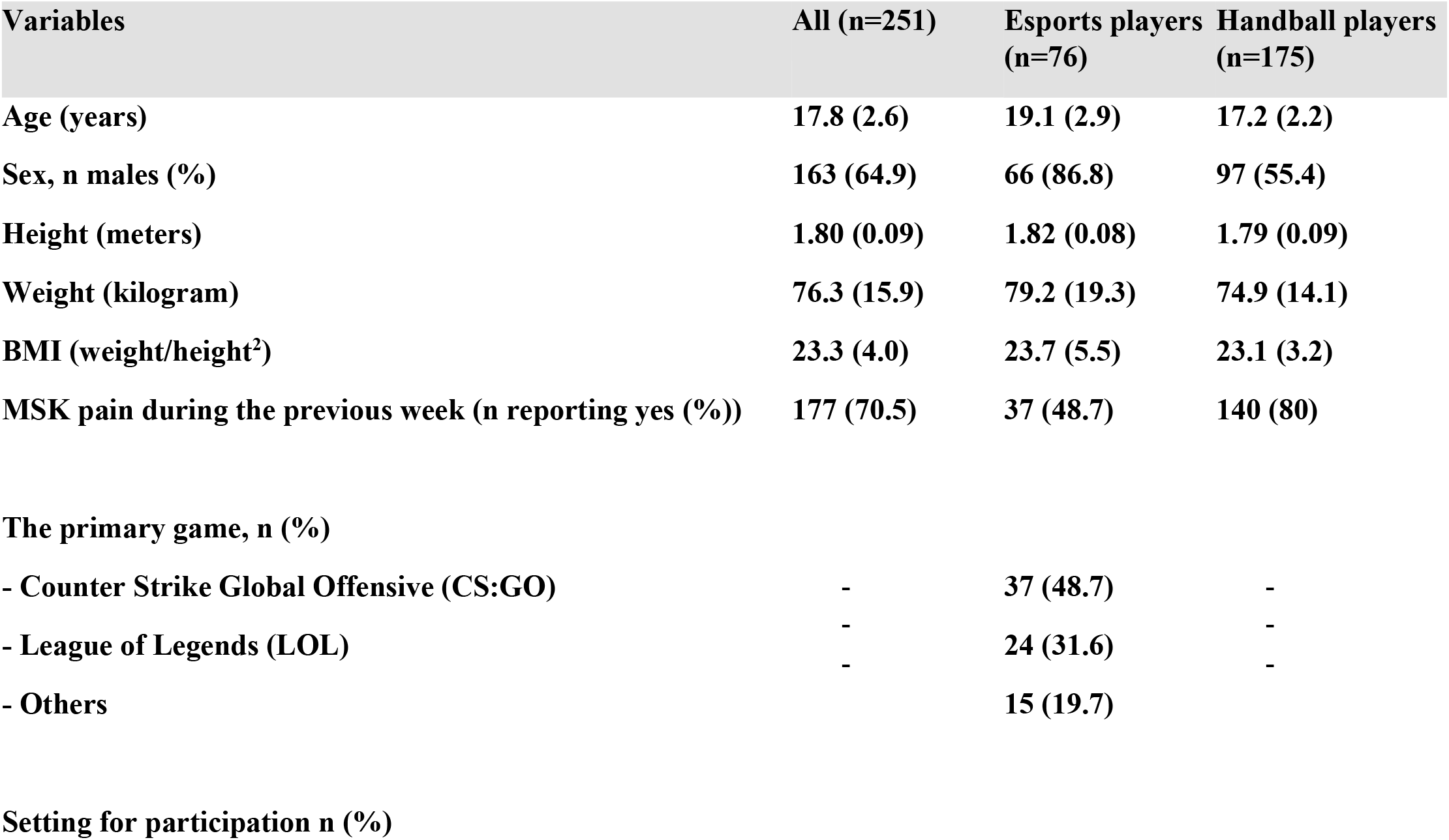
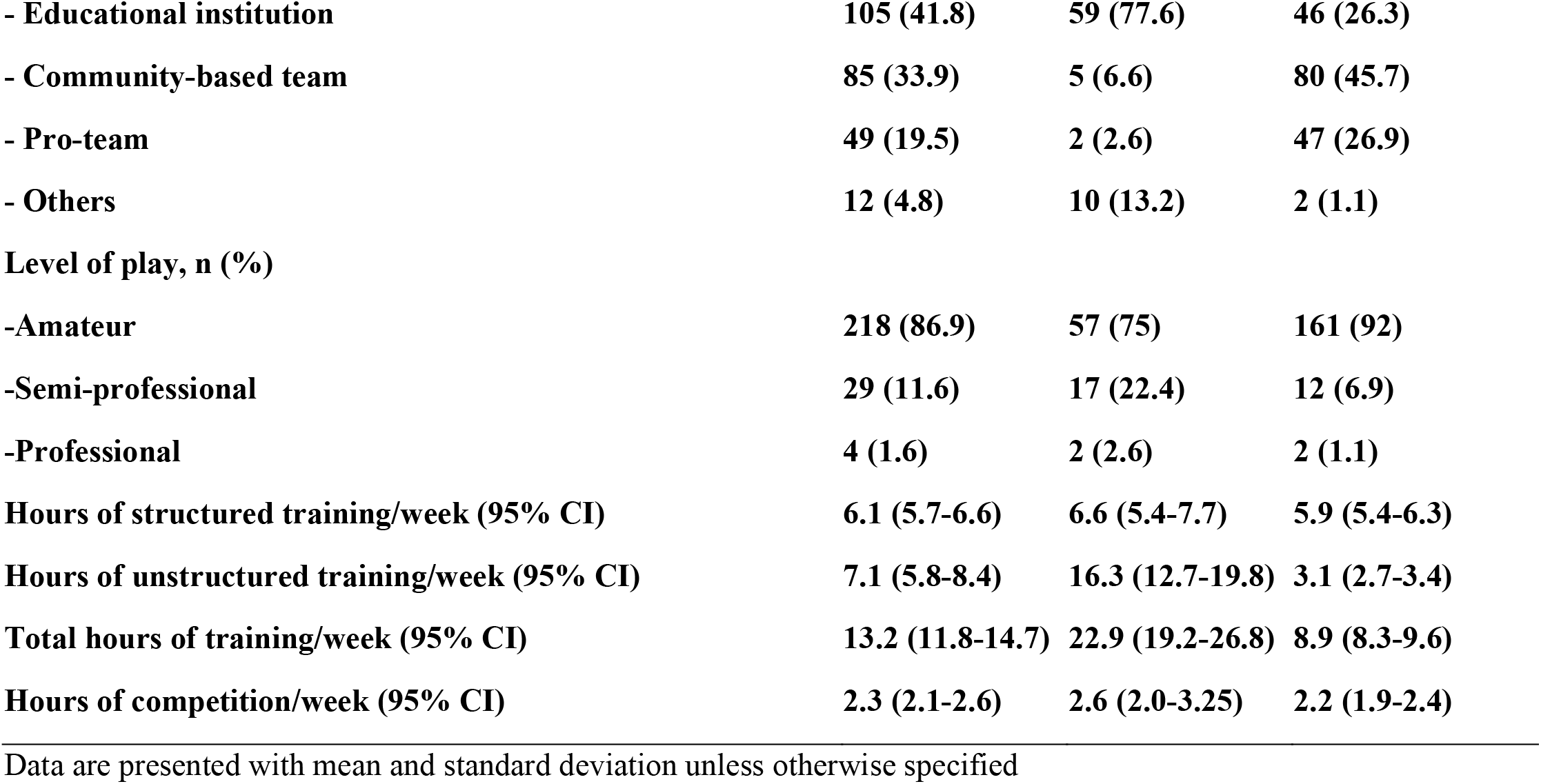
Characteristics of the 251 players (76 esport players and 175 handball players).

| Variables | All (n=251) | Esports players (n=76) | Handball players (n=175) |
| --- | --- | --- | --- |
| Age (years) | 17.8 (2.6) | 19.1 (2.9) | 17.2 (2.2) |
| Sex, n males (%) | 163 (64.9) | 66 (86.8) | 97 (55.4) |
| Height (meters) | 1.80 (0.09) | 1.82 (0.08) | 1.79 (0.09) |
| Weight (kilogram) | 76.3 (15.9) | 79.2 (19.3) | 74.9 (14.1) |
| BMI (weight/height<sup>2</sup>) | 23.3 (4.0) | 23.7 (5.5) | 23.1 (3.2) |
| MSK pain during the previous week (n reporting yes (%)) | 177 (70.5) | 37 (48.7) | 140 (80) |
| The primary game, n (%) |  |  |  |
| - Counter Strike Global Offensive (CS:GO) | - | 37 (48.7) | - |
| - League of Legends (LOL) | - | 24 (31.6) | - |
| - Others | - | 15 (19.7) | - |
| Setting for participation n (%) |  |  |  |
| - Educational institution | 105 (41.8) | 59 (77.6) | 46 (26.3) |
| - Community-based team | 85 (33.9) | 5 (6.6) | 80 (45.7) |
| - Pro-team | 49 (19.5) | 2 (2.6) | 47 (26.9) |
| - Others | 12 (4.8) | 10 (13.2) | 2 (1.1) |
| **Level of play, n (%)** |  |  |  |
| -Amateur | 218 (86.9) | 57 (75) | 161 (92) |
| -Semi-professional | 29 (11.6) | 17 (22.4) | 12 (6.9) |
| -Professional | 4 (1.6) | 2 (2.6) | 2 (1.1) |
| Hours of structured training/week (95% CI) | 6.1 (5.7-6.6) | 6.6 (5.4-7.7) | 5.9 (5.4-6.3) |
| Hours of unstructured training/week (95% CI) | 7.1 (5.8-8.4) | 16.3 (12.7-19.8) | 3.1 (2.7-3.4) |
| Total hours of training/week (95% CI) | 13.2 (11.8-14.7) | 22.9 (19.2-26.8) | 8.9 (8.3-9.6) |
| Hours of competition/week (95% CI) | 2.3 (2.1-2.6) | 2.6 (2.0-3.25) | 2.2 (1.9-2.4) |

Data are presented with mean and standard deviation unless otherwise specified

**Table 2.** Use of analgesics, physical activity levels, activity-limiting burnout, and sleep patterns.

| Variables * | All (n=249) | Esports players (n=76) | Handball players (n=173) |
| --- | --- | --- | --- |
| Analgesics utilization for pain during the previous 3 months (n reporting yes (%)) | 92 (36.9) | 26 (34.2) | 66 (38.2) |
| **Frequency of analgesics utilization during the previous 3 months, n (%)** |  |  |  |
| - Daily | 6 (6.5) | 3 (11.5) | 3 (4.5) |
| - Several times per week | 9 (9.8) | 1 (3.8) | 8 (12.1) |
| - Weekly | 26 (28.3) | 8 (30.8) | 18 (27.3) |
| - Monthly | 23 (25.0) | 4 (15.4) | 19 (28.8) |
| - Seldom | 28 (30.4) | 10 (38.5) | 18 (27.3) |
| **Variables §** | **All (n=247)** | **Esports players (n=76)** | **Handball players (n=171)** |
| **Physical activity levels** |  |  |  |
| **N days/week of physical activity at moderate intensity** |  |  |  |
| - 0 days | 10 (4.0) | 8 (10.5) | 2 (1.2) |
| - 1 day | 18 (7.3) | 14 (18.4) | 4 (2.3) |
| - 2 days | 48 (19.4) | 10 (13.2) | 38 (22.2) |
| - 3 days | 44 (17.8) | 14 (18.4) | 30 (17.5) |
| - 4 days | 37 (15.0) | 11 (14.5) | 26 (15.2) |
| - 5 days | 44 (17.8) | 10 (13.2) | 34 (19.9) |
| - 6 days | 18 (7.3) | 4 (5.3) | 14 (8.2) |
| - 7 days | 28 (11.3) | 5 (6.6) | 23 (13.5) |
| **N days/week of physical activity at high intensity** |  |  |  |
| - 0 days | 23 (9.3) | 18 (23.7) | 5 (2.9) |
| - 1 day | 35 (14.2) | 15 (19.7) | 20 (11.7) |
| - 2 days | 50 (20.2) | 21 (27.6) | 29 (17.0) |
| - 3 days | 50 (20.2) | 8 (10.5) | 42 (24.6) |
| - 4 days | 39 (15.8) | 6 (7.9) | 33 (19.3) |
| - 5 days | 36 (14.6) | 5 (6.6) | 31 (18.1) |
| - 6 days | 13 (5.3) | 2 (2.6) | 11 (6.4) |
| - 7 days | 1 (0.4) | 1 (1.3) | 0 (0.0) |
| **Activity-limiting burnout during the previous 3 months n (%)** | **89 (30)** | **26 (34)** | **63 (37)** |
| **Sleep** |  |  |  |
| **Hours of sleep during the night (95%CI)** | **7.5 (7.1-7.9)** | **7.7 (6.4-9.0)** | **7.4 (7.3-7.6)** |
| **Trouble falling asleep % (95%CI)** |  |  |  |
| - Yes, most nights | 13.4 (8.9-17.8) | 17.1 (9.2-26.3) | 11.7 (7.0-16.4) |
| - Yes, some nights | 46.2 (40.1-52.6) | 40.8 (30.3-51.3) | 48.5 (40.4-56.1) |
| - No, not at all | 38.9 (32.8-44.9) | 40.8 (28.8-51.3) | 38.0 (30.4-45.6) |
| - Don't know | 1.6 (0.4-3.2) | 1.3 (0.0-3.9) | 1.8 (0.0-4.1) |
| **Waking several times/nights % (95%CI)** |  |  |  |
| - Yes, most nights | 6.1 (3.2-9.3) | 10.5 (3.9-18.4) | 4.1 (1.2-7.6) |
| - Yes, some nights | 36 (29.6-42.1) | 38.2 (27.6-50.0) | 35.1 (28.1-42.1) |
| - No, not at all | 55.9 (49.8-61.3) | 51.3 (39.5-63.1) | 57.9 (50.3-65.5) |
| - Don't know | 2.0 (0.4-4.0) | 0.0 (0.0-0.0) | 2.9 (0.6-5.8) |
| **Trouble sleeping through the night % (95%CI)** |  |  |  |
| - Yes, most nights | 6.9 (4.0-10.1) | 9.2 (3.9-17.1) | 5.8 (2.9-9.9) |
| - Yes, some nights | 30.8 (25.5-36.4) | 27.6 (18.4-39.5) | 32.2 (25.1-39.2) |
| - No, not at all | 60.3 (53.9-66.4) | 63.2 (51.3-73.7) | 59.1 (51.5-66.7) |
| - Don't know | 2.0 (0.4-4.0) | 0.0 (0.0-0.0) | 2.9 (0.6-5.8) |
| **Waking feeling tired % (95%CI)** |  |  |  |
| - Yes, most mornings | 51.8 (45.7-57.5) | 48.7 (36.8-60.5) | 53.2 (45.6-60.2) |
| - Yes, some mornings | 40.9 (35.2-46.6) | 43.4 (31.6-55.3) | 39.8 (32.7-46.8) |
| - No, not at all | 5.3 (2.4-8.1) | 5.3 (1.3-10.5) | 5.3 (2.3-8.8) |
| - Don't know | 2.0 (0.4-3.6) | 2.6 (0.0-6.6) | 1.8 (0.0-4.1) |

Data is presented with absolute (n) and percentage values (%) unless otherwise specified. \* Data available for 76 esports players and 173 handball players (249 in total). § Data are available for 76 esports players and 171 handball players (247 in total).

### MSK pain prevalence, location, and association with the type of activity

The primary analysis showed that esports players were significantly less likely to experience MSK pain during the previous week compared to handball players (OR 0.24, 95%CI [0.13-0.43], Chi^2^ p-value > 0.001). Thirty-seven (48.7%) esports players and 140 (80%) handball players experienced MSK pain during the previous week. Of these, the most common pain sites were the back (32%) and the hand/fingers/wrist (24%) among esport players and the knees (26%) and shoulders (13%) among handball players. Of those reporting pain, 54% of the esport players and 52% of the handball players experienced pain daily or several times per week. Among those reporting pain, 16% of the esports players and 30% of the handball experienced activity-limiting pain. On average, esports players and handball players reported worst pain intensities during the previous week of 4.4 (95%CI [3.6-5.1]) and 4.9 (95%CI [4.7-5.3]), respectively.

### Association between MSK pain and training volume

We found no significant difference in total weekly training volume among esports players with MSK pain (22.3 hours/week, 95%CI [17.1-27.45]) and without MSK pain (23.6 hours/week, 95%CI [17.8-29.4]) during the previous week (p-value = 0.727). Further, we found no significant difference in total weekly training volume among handball plays with MSK pain (9.2 hours/week, 95%CI [8.5-9.9]) and without MSK pain (8.1 hours/week, 95%CI [6.8-9.3]) during the previous week (p-value = 0.128).

### Explorative analysis

Due to a substantial difference in weekly training volume between two groups we conducted an explorative test for comparison. From the analysis we found that the difference was statistically significant (p-value < 0.001) with the esports players having 13.2 hours of additional training volume per week compared to the handball players. The difference remained statistically significant when hours of weekly competition or tournament activities were included in the analysis (14.4 hours/week, 95%CI [10.4-18.5], p-value < 0.001).

## DISCUSSION

### Summary of findings

As hypothesised, we found that the prevalence of MSK pain during the previous week was higher among handball players than esports players and the likelihood of experiencing MSK pain was significantly lower among esports players. Although the prevalence of weekly MSK pain was high, not all esport players nor handball players were limited during their activity due to pain. Furthermore, we found no significant difference in training volume among players with and without MSK pain within neither esports nor handball. The prevalence of activity-limiting pain was lower among esports players compared to handball players. In both groups, more than 1 in every 3 had experienced activity-limiting burnout during the previous 3 months.

### Comparison with previous findings

To the best of our knowledge, this is the first study to compare MSK pain prevalence between esports and another type of activity appealing to individuals in the same age group. Previous studies on MSK pain and health in esports are limited by not including a control group of non-esports players for comparison (1,6). Therefore, it has not been possible to conclude if young esports players are at greater risk of experiencing MSK pain compared to young individuals participating in other activities.

Findings on MSK pain prevalence, pain location and cases of activity-limiting pain among esports players from the current study are similar to findings from previous studies among amateur and professional esports players competing in computer-based games (1,6). As such, there is consistent evidence to suggest that MSK pain is indeed prevalent in esports with more than 4 in every 10 players reporting pain. In addition, more than 50% of the esport players and handball players affected by pain, experienced pain daily or several times per week. This is especially interesting from an esports perspective as esports is characterised as a non-contact-activity. These findings underline the need for more knowledge on the mechanisms and triggers for MSK pain in esports.

Contrary to previous findings, we found no association between training volume and MSK pain among esports players in the current study (1). As such, we were unable to replicate the results from the study by Lindberg et al (1). These inconsistent findings warrant further investigations regarding this possible association. Interestingly, we found more than 1 in every 3 players from both esports and handball experienced activity-limiting burnout during the previous 3 months. These findings support prior speculation of activity-limiting burnout being prevalent in esport. A concern which is also being addressed by player-organisations (12). However, our findings suggest that activity-limiting burnout is similarly prevalent in handball. This despite esports players practiced their activity for significantly more hours per week compared to handball players. As such, this issue may be related to other factors than type of activity and training volume. An association between training volume and activity-limiting burnout has been suggested previously (7) however, due to the small sample size and lack of a prior hypothesis, this relationship was not explored further in the current study.

### Limitations

First, the number of included esports players was smaller than expected. From engaging with the esport community, this is likely related to an intensified competition to engage Danish esports players in research projects. Second, due to the study design, it remains unknown if the MSK pain was initiated via either esports or handball activities or not. As stated by both clinicians and research, there is a need for prospective studies investigating the association between esports activity and MSK pain prevalence (1,4). Thirdly, physical activity levels were not assessed with a validated measure in the current study. In previous studies, we tried to assess physical activity levels with both the International Physical Activity questionnaire short form and the Physical Activity Scale 2 with little success. From our experience, it seems these measures are inadequate to capture physical activity levels in populations of young esports players. The reason for this is unknown but may be related to comprehensibility, lengths, and complexity of the measures. Nevertheless, an objective quantitative measure of physical activity (e.g., use of ActiGraphs or similar wearables) would have been preferred as self-reported measures of physical activity may be susceptive to bias (15). Lastly, the underlying mechanisms for MSK pain are likely to differ between esports and handball. As such, it may be assumed that the mechanism for MSK pain in esport might be more comparable to other non-contact activities such as dart, billiards, or pétanque. However, the age of individuals from Denmark engaged in these activities are considerably older compared to those engaged with esports (8). A similar issue may arise when comparing MSK pain between young esports players and adult office workers. As such, the difference in age is likely to distort any comparison between esports and these activities as the prevalence of common MSK pain conditions such as low back pain increases with age (16). Therefore, comparing MSK pain prevalence between two activities that appeal to the same age group is likely to be a more appropriate comparison.

### Implication of findings

It is widely acknowledged that participating in physical activities, such as handball, is positively associated with good physical health (17). However, like several other activities, handball is associated with health risks such as pain and injuries (18). Although the findings from the current study suggest the risk of MSK pain in esports is significantly lower when compared to handball, we also found an extensive training volume in esports. Compared to the training volume among handball players, the training volume of esports players is often associated with physical inactivity. This is important as prolonged physical inactivity is associated with chronic diseases such as cancer, diabetes, and cardiovascular conditions (19). As such, future research should investigate which type of physical activity appeals to esports players and develop methods to implement physical activity into esports.

## CONCLUSION

These findings suggest that young esports players are not at increased health risk in terms of experiencing MSK pain compared to young players participating in handball. This despite esports players practiced their activity for additional 12 hours per week compared to handball players. More than 1 in every 3 had experienced activity-limiting burnout in both esports and handball despite a significant difference in weekly training volume. This indicate that other factors besides type of activity and training volume may influence the occurrence of activity-limiting burnout.

## Data Availability

The authors did not obtain informed consent from the included participants to share data with a third party. Therefore, data from the current study cannot be shared.

